# Molecular landscapes of oral cancers of unknown etiology

**DOI:** 10.1101/2023.12.15.23299866

**Authors:** Sophie Deneuve, Béatrice Fervers, Sergey Senkin, Liacine Bouaoun, Olivia Pérol, Bérénice Chavanel, Lingeng Lu, Isabelle Coste, Toufic Renno, Jiri Zavadil, François Virard

**Affiliations:** Department of Otolaryngology-Head and Neck Surgery, CHU Rouen, Rouen, France; QuantIF-LITIS Team EA4108, University of Rouen, Rouen, France; Centre Léon Bérard, Département Prévention Cancer Environnement, INSERM U1296 Unit Radiation: Defense, Health Environment, Lyon, France; International Agency for Research on Cancer WHO, Genomic Epidemiology Branch, Lyon, France; International Agency for Research on Cancer WHO, Environment and Lifestyle Epidemiology Branch, Lyon, France; International Agency for Research on Cancer WHO, Epigenomics and Mechanisms Branch, Lyon, France; Department of Chronic Disease Epidemiology, Yale School of Public Health, School of Medicine, New Haven, CT 06520, USA; Center for Biomedical Data Science, Yale University, 60 College Street, New Haven, CT 06520, USA; Yale Cancer Center, Yale University, 60 College Street, New Haven, CT 06520, USA; University Claude Bernard Lyon 1, INSERM U1052–CNRS UMR5286, Cancer Research Center, Centre Léon Bérard, Lyon, France; University of Lyon, Faculty of Odontology, Hospices Civils de Lyon, Lyon, France

**Keywords:** Squamous cell carcinoma of head and neck, early onset mobile tongue cancer, cancer risk factors, mutational signature, cancer driver gene, transcriptome

## Abstract

The incidence of the mobile tongue cancer in young patients has been rising. This oral cancer (OC) type has no identified risk factors (NIRF), no established molecular markers and is not yet recognized as a distinct clinical entity.

To understand this emerging malignancy, we innovatively analyzed the public head and neck cancer multi-omics data. We identified mutational signatures that successfully stratified 307 OC and 109 laryngeal cancer cases according to their clinico-pathological characteristics. The NIRF OCs exhibited significantly increased activities of endogenous clock-like and APOBEC-associated mutagenesis, alongside specific cancer driver gene mutations, distinct methylome patterns and prominent antimicrobial transcriptomic responses. Furthermore, we show that mutational signature SBS16 in OCs reflects the combined effects of alcohol drinking and tobacco smoking.

Our study characterizes the unique disease histories and molecular programs of the NIRF OCs revealing that this emerging cancer subtype is likely driven by increased endogenous mutagenesis correlated with responses to microbial insults.

## Introduction

Head and neck squamous cell carcinoma (HNSCC) is responsible for over 800,000 new cancer cases and 450,000 deaths annually, per the Global Cancer Observatory estimates ^1^. HNSCC’s originate from the epithelium lining of the upper aero-digestive tract of the oral cavity (OC), oropharynx, hypopharynx, and larynx (LX), with tobacco use and alcohol consumption, as well as infection with human papilloma virus (HPV) being the major risk factors ^2^. The particular clinical and histological presentations and better prognosis of HPV-positive cancers of the oropharynx have prompted the consideration of two distinct HNSCC subtypes: a HPV-positive HNSCC, mainly at the oropharyngeal subsite, and the conventional group of HPV-negative HNSCC at different anatomical sites associated with exposure to tobacco-derived carcinogens, excessive alcohol consumption, or both ^3^.

In addition to the growing incidence of HPV-related oropharyngeal SCC, a globally increasing incidence of OC cancers has been observed, involving non-smoking and non-drinking patients with no considerable role for HPV infection ^4–7^. In the United States, both the Global Cancer Observatory and Surveillance, Epidemiology, and End Results (SEER) database report a rising incidence of oral tongue squamous cell carcinomas (Supplemental Figure S1), especially among young adults without any history of tobacco smoking or alcohol drinking ^8–10^. Moreover, an increased incidence of SCC of the gingiva and the hard palate has been reported among older patients, mostly women, with no history of tobacco or alcohol use. ^11^

Consequently, there is a need for better understanding of the etiology factors as well as molecular programs associated with tumors characterized by the OC site-specific occurrence and the absence of known risk factors (hereafter referred to as NIRF, for No Identified Risk Factors). Detailed molecular characterization of the NIRF OC cancers can help identify the underlying molecular mechanisms as well as refine the histopathological classification of OC cancers with possible implications for improved diagnosis and treatment.

Cancer genome mutation landscapes result from an imbalance between DNA damage by exogenous or endogenous factors and the DNA repair system efficiency ^12^. The mutagenic process-specific patterns, so called mutational signatures, are determined by a blind source separation-based mathematical process deconvolving mixed mutation spectra ^13–15^. Comprehensive identification of mutational signatures can thus reveal the DNA-damaging processes operative during a cancer’s history. Previous reports on various HNSCC cohorts aimed to decipher various mutational processes, identifying the clock-like mutagenesis (COSMIC signatures SBS1 and SBS5) ^14,16–20^, mutagenesis by APOBEC enzymes (SBS2, SBS13) ^14,16–21^, tobacco smoking and putative alcohol exposure (SBS4, SBS16) ^14,17–20^, DNA repair deficiency (SBS3, SBS6) ^14,16–18^, DNA damage by reactive oxygen species (SBS18) ^14,19,20^, and various mutational signatures of unknown origin (SBS17, SBS33, SBS40) ^14,20^. However, most of these studies did not associate the reported mutational signatures with specific HNSCC subsites ^14,18,21,22^. However, Alexandrov et al. and South et al. reported differences between cancers of the LX and OC with regards to the tobacco smoking signature (SBS4) ^19,20^. Conversely, studies focusing on specific subsites made no distinction between the candidate carcinogen exposures ^16^. Some reports noted varying rates of the endogenous signature SBS5 between smokers and non-smokers, yet none have identified mutational processes operating specifically in the NIRF OC cancer subgroup ^19,20,23,24^. With the exception of the APOBEC mutational signature reported to correlate with the HPV infection ^17,21,22^, none of the signatures identified to-date have been associated with distinct clinical presentations or specific cancer development trajectories.

Using innovative analysis of complex molecular cancer data from public sources, here we aimed to characterize relevant mutational processes and driver gene alterations underlying distinct trajectories of HPV-unrelated HNSCC development ^25^. We particularly focused on the molecular characterization of the NIRF patients, including early onset tongue cancer cases and older patients with OC cancer from non-tongue subsites. We next aimed to establish clinically relevant molecular stratification of the studied HNSCCs (involving OC and LX only) according to the most active mutational processes further supported by specific correlative epigenetic and transcriptomic programs. We next investigated novel aspects of exposure- and anatomical site-specific molecular programs related to alcohol drinking and tobacco smoking. The NIRF OC cancer groups were further characterized in terms of specific biological processes, suggesting the bases for their development. Overall, our study provides a new conceptual basis for future molecular epidemiology investigations of the NIRF OC cancers that are of emerging concern due to unresolved etiology and globally increasing incidence.

## Results

### Patient clustering based on *de novo* mutational signatures

To understand the possible origins and specific molecular features of the NIRF OCs, we performed an in-depth analysis of mutational signatures for non-HPV related HNSCC, ie OC and LX squamous cell carcinoma (SCC).

Mutational signature extraction was conducted on 425 HNSCC samples involving 314 (74%) OC and 111 (26%) LX SCC from The Cancer Genome Atlas (TCGA). After initial sample filtering (see Methods and Supplementary Figure S2), we performed mutational signature analyses of the 347 remaining samples using the 384-channel (i.e. transcription strand-biased) signature versions that ensure a higher sensitivity. We identified four distinct *de novo* signatures SBS384A-C (Figure 1a) closely matching several known COSMIC signatures, as follows: SBS384A - SBS4 (tobacco smoking exposure); SBS384B - SBS2 and SBS13 (APOBEC enzyme mutagenesis); SBS384C - SBS1 (5-methyl-cytosine deamination), and SBS384D - a set of mutually similar signatures SBS5 (clock-like, unknown etiology), SBS92 (new tobacco smoking signature) and SBS16 (unknown etiology, putatively associated with alcohol consumption) (Supplementary Figure S3).

**Figure 1:**
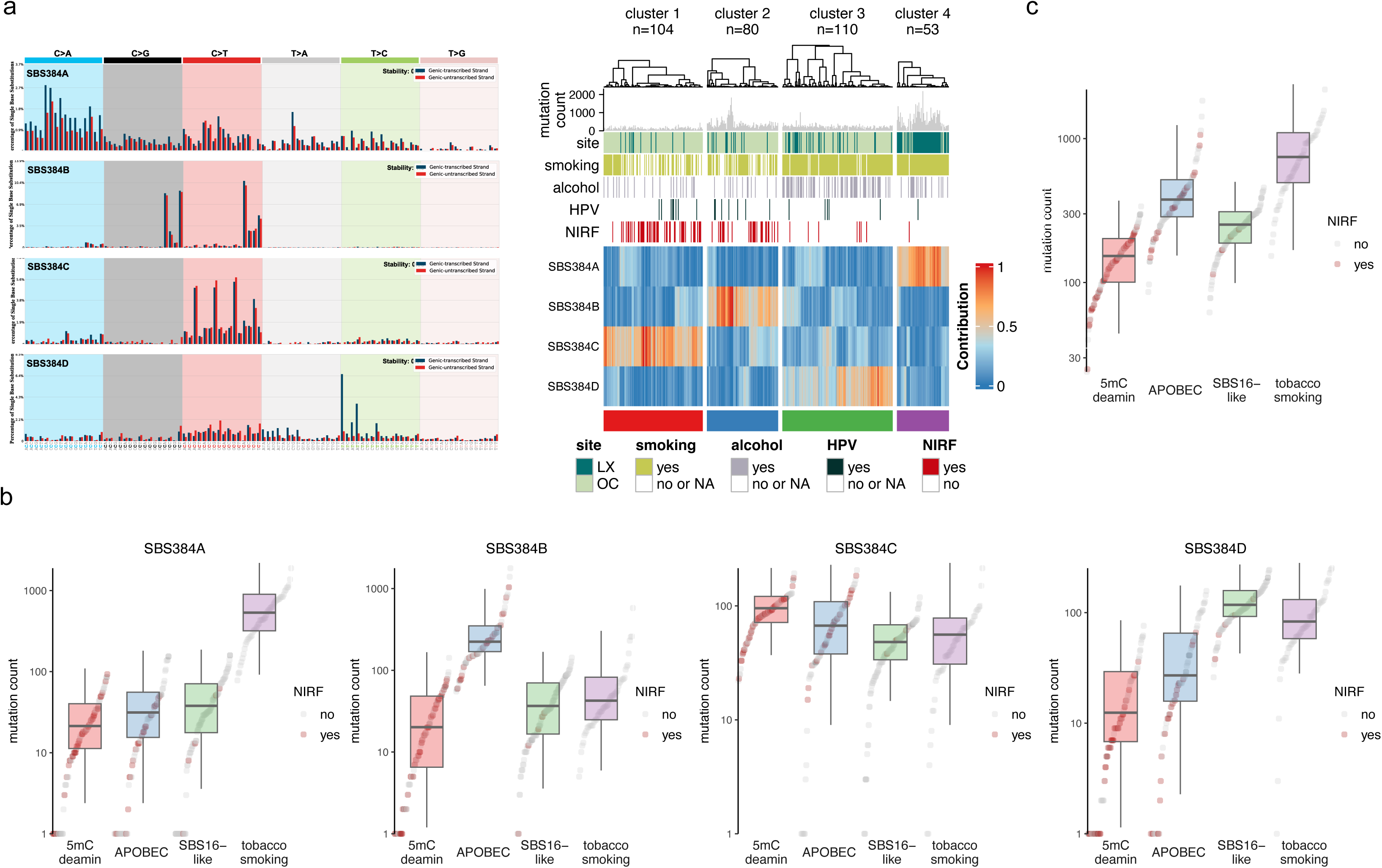
Mutational signatures that characterize the major OCSCC and LXSCC subsets. (a) *De novo* transcriptional strand-asymmetry signatures (left panels, signatures SBS384A through SBS384D extracted by NMF, see Methods for details) determine four major patient clusters (heatmap, right panel). The heatmap is based on relative per-sample contribution of each mutational signature. Heatmap annotation tracks and color legends: Mutation count histogram, anatomical subsites (LX = larynx, OC = oral cavity), tobacco smoking status (NS = non-smoker, S = smoker), HPV infection status and no identified risk factors (NIRF). (b) Per-sample and per-cluster distributions of the total mutation counts. (c) Absolute mutation counts attributed to each *de novo* signature in each cluster. Cluster names reflect the matching putative mutagenic process/COSMIC signature with the highest contribution in each cluster (5mC deamin = spontaneous deamination of methylated cytosine/COSMIC SBS1).

Based on the relative per-sample contribution of the four *de novo* signatures, the samples were grouped into four distinct patient/signature-based clusters (Figure 1a). In each cluster, the absolute mutation counts were dominated by the activity of one *de novo* signature (Figure 1b), with more low-mutation burden samples in Clusters 1 (median=139, range=24-338) and 3 (median=230, range=62-457) and high mutation-count samples in Clusters 2 (median=344, range=86-1835) and 4 (median=682, range=153-2193) (Figure 1c). The high relative enrichment of each *de novo* mutational signature (Figure 1a) reflected well the absolute counts of signature-attributed mutations (Figure 1b). Specifically, signature SBS384C (putative SBS1) appeared to operate at the highest levels in Cluster 1 (renamed 5mC deamin cluster, n=104) (Figure 1b). The mutation counts supported the overall high relative enrichment of SBS384B (putative APOBEC, n=80) in Cluster 2 (renamed APOBEC cluster). SBS384D (resembling COSMIC signatures SBS16, SBS5, or SBS92) was enriched in Cluster 3 (renamed SBS16-like, n=110). Cluster 4 (renamed tobacco smoking cluster, n=53) was dominated by the SBS384A signature (putative SBS4) and by the associated high mutation counts.

### Clinico-pathological characteristics match the patient-signature clusters

The NIRF cancer patients were more often found in the 5mC deamin and APOBEC clusters (61% or 32% of 72 NIRF cases, respectively) (Figure 2a). The 5mC deamin cluster further contained the majority of the young patients (< 40 years; 80%, or 8/10), while most of the 91 elderly patients (≥70 years) were equally distributed between the 5mC deamin and APOBEC and clusters (37%, or 34/91) and 35%, or 32/91), respectively)(Figure 2b). The SB16-like and the tobacco smoking clusters included the majority of drinkers (77.8 %, or 14/18 and 91.1 %, or 51/56, respectively) and smokers (100 %, or 53/53 and 95.4 %, or 103/108, respectively) (Figure 2c and 2d). We further observed a higher proportion of female patients in both the APOBEC and 5mC deamin clusters whereas males were more represented in the tobacco-smoking and SBS16-like clusters (Figure 2e).

**Figure 2:**
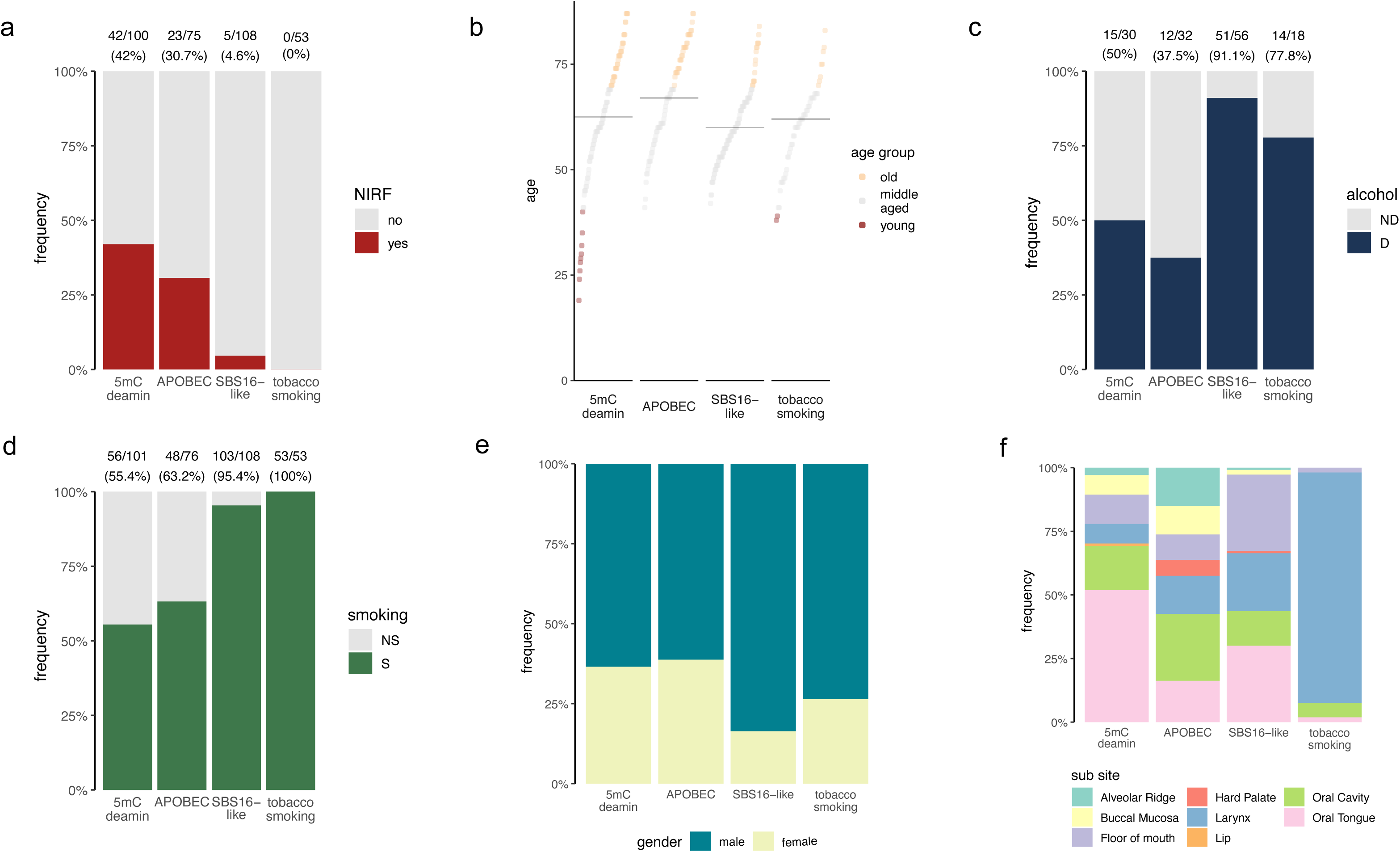
Mutational signature cluster-based demographic and clinical presentation summaries. (a) Proportion of the NIRF cases across clusters. (b) Distribution of patients by age (young patients ≤40; 40<middle aged<70; old ≥70) per cluster. (c) Relative alcohol consumption and (d) tobacco smoking status distributions across clusters (ND = non-drinker, D = drinker, NS = non-smoker, S = smoker). (e) Relative anatomical sub-site distribution in each cluster, for all analyzed patients. (f) Relative gender-based patient distribution in each cluster. NA status data are not plotted.

The cancer anatomical subsite distribution also tended to follow the clusters (Figure 2f and Supplementary Figure 5). A half (52%, or 49/95) of the cancer originating from the larynx were in the tobacco smoking cluster while tumors affecting the floor of the mouth were enriched in the SBS16-like cluster (63% = 33/54). Moreover, more than half of tumors involving the oral tongue were in the 5mC deamin cluster (54% = 55/103), while cancer subsites involving the hard palate (83 % = 5/6) or the alveolar ridge (75% = 12/16) were found mainly in the APOBEC cluster. Finally, tumors affecting the buccal mucosa were detected in the two NIRF enriched clusters, 5mC deamin (42%) = 8/19) and APOBEC (9 of 19, or 47%).

Collectively, these data show that mutational signature-based clustering recapitulates the clinico-pathological characteristics of patients with or without identified risk factor and supports the potential clinical relevance of the patient stratification according to the *de novo* mutational signature contents.

### COSMIC mutational signatures operating in OC and LX SCC

To further characterize the individual *de novo* signatures, which can often represent mixed patterns, we performed their further decomposition into the COSMIC reference signatures. This process, performed by using SigProfilerExtractor (SPE), determined 8 COSMIC signatures: SBS1, SBS2, SBS4, SBS5, SBS6, SBS7a/b, SBS13 and SBS16. COSMIC signature SBS4 (tobacco smoking) constituted 60% of the SBS384A *de novo* signature. Nearly 90% of SBS384B (cosine similarity = 0.989) was jointly generated by COSMIC signatures SBS2/SBS13 (mutagenesis by APOBEC enzymes). SBS384D consisted of a combination of three distinct mutational signatures of approximately equal contribution: COSMIC SBS1, resulting from methylcytosine deamination; SBS6, a mutational process putatively linked to defective DNA mismatch repair, and SBS5, a clock-like signature of unknown origin. A low contribution of SBS7a and SBS7b, linked to UV exposure, was observed in SBS384C (8% and 7%, respectively). Finally, SBS384D was mainly the result of SBS5 (70%), with a contribution of SBS16 (20%), a signature putatively linked to alcohol exposure (Supplemental Figure S6a).

### SBS5 can mask mutational signatures associated with exogenous exposures

Sample clustering based on the COSMIC signature decomposition and attribution did not reproduce the sample clustering based on the *de novo* signature contents (Figure 3a, and Supplemental Figure S6b). This was mainly due to the activity broadly attributed to signature SBS5 (such as its 29.06 % attribution in SBS384C) (Supplemental Figure S6a). The SPE tool assumes that SBS1 and SBS5 are always present and active in all tissues ^26^. We hypothesized that this could overestimate the respective contributions of SBS1 and SBS5 in the analyzed data set. To test this hypothesis, we performed a signature analysis complementary to the SPE decomposition, by using the Mutational Signature Analysis (MSA) ^27^ attribution approach (see Methods). Using the signature set proposed by the SPE decomposition (SBS1, SBS2, SBS4, SBS5, SBS6, SBS7a/b, SBS13 and SBS16), we observed an overall reduced attribution of SBS5 resulting in restored stability of patient clusters (Figure 3a, Supplementary Figure S6c). Next, we applied the MSA-based attribution using the entire COSMIC signature set (70 signatures). This approach resulted in cluster stability consistent with the *de novo* signature-based clustering (Figure 3a, Supplementary Figures S7a,b), and the main contributions of each principal mutational process to a given patient cluster were restored (Figure 3b, Supplementary Figure S7c). Thus, the overfitting of the SBS5 signature within the SPE decomposition step due to the default inclusion of the signature can be effectively resolved by the MSA-based optimized refitting approach.

**Figure 3:**
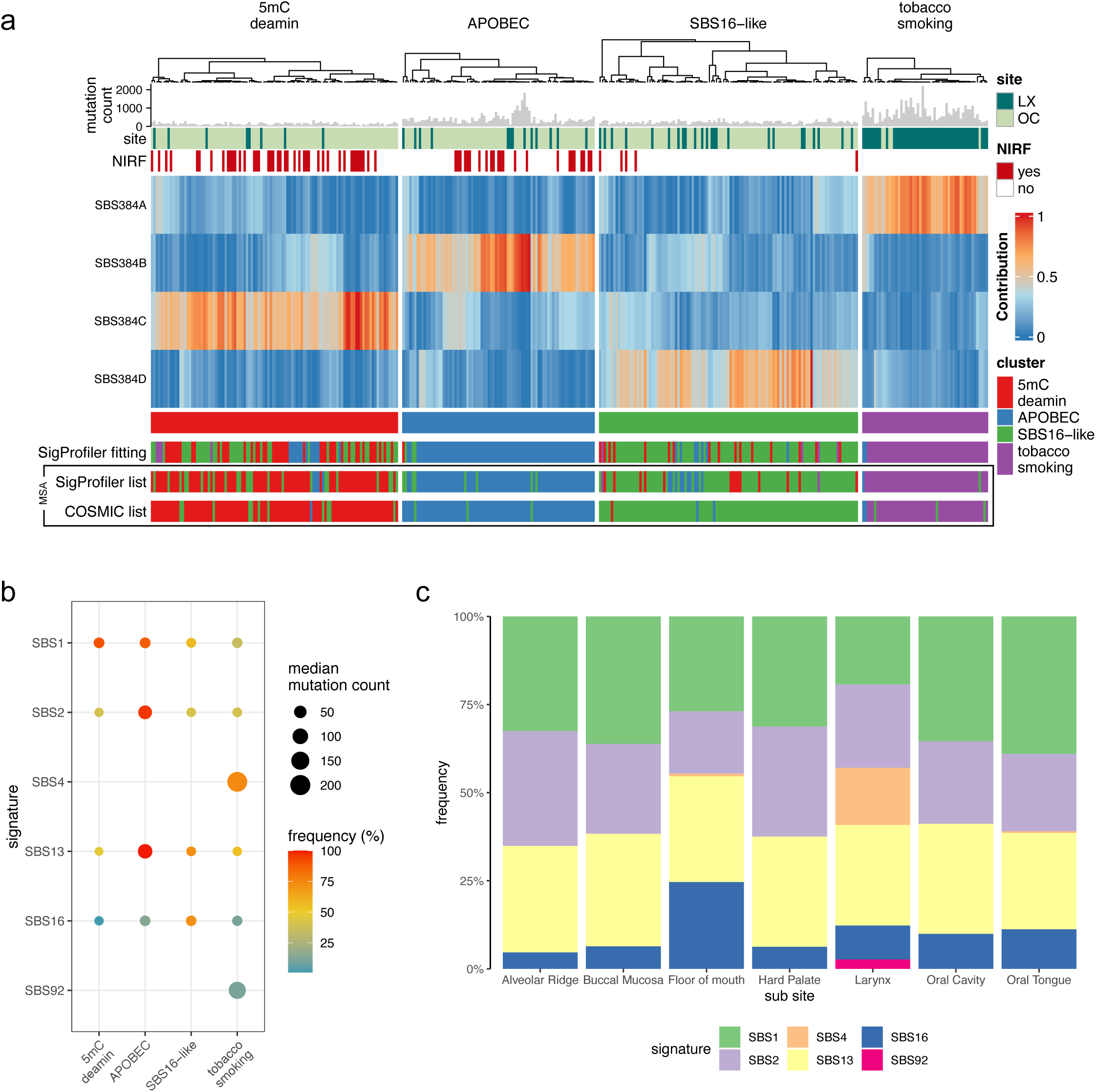
Attribution of *de novo* mutational signatures to COSMIC signatures by MSA fitting. (a) Heatmap showing *de novo* mutational signatures across patients that restore the original clusters described in Fig. 1a. Heatmap tracks show the mutation count histogram, anatomical subsites (LX = larynx, OC = oral cavity), no identified risk factor status (NIRF). Cluster color labels under the heatmap reveal the comparative sample redistribution outcomes after SigProfiler-based fitting and MSA fitting. The MSA-based fitting was performed by using the same signature set identified by SigProfiler fitting, or with a signature set encompassing all COSMIC signatures (v3.2). (b) Bubble plot documenting the predominant, expected or newly revealed signatures in each cluster, with median mutation count attributed to each signature (bubble size) and positive sample frequency (bubble color). (c) Relative distribution of mutational signatures listed in (b) according to the specific head-and-neck anatomical subsites.

The possible masking of other signatures by SBS5 was based on several lines of evidence. Firstly, SBS5 in its transcriptional strand bias version, is highly similar with signatures SBS16 and SBS92 (Supplementary Figure S3). Indeed, MSA analysis could resolve the broad SPE-suggested presence of SBS5 into biologically plausible assignment of a newly described tobacco-smoking-related signature SBS92 almost exclusively to the LX tumors in the SBS4 dominated tobacco-smoking cluster (Figure 3b and Supplementary Figure S7a,c). Secondly, the resolution of SBS5 revealed an increased presence of SBS16 in smokers who were drinkers without compromising the stability of the *de novo* signature-based clusters (Supplementary Figures S7 and S8).

### Tobacco smoking induces varying, site-specific mutational processes

Interestingly, by using the NNLS-based MSA method of signature fitting, signature SBS4 related to tobacco smoking exposure (particularly to the effects of polycyclic aromatic hydrocarbons such as benzo[*a*]pyrene ^20,28–30)^ was detected in tumors in the tobacco smoking cluster (involving mostly the LX) but not in the SBS16-like cluster (involving mostly the OC) (Figures 3b, 4 and Supplemental Figure S7a,c) even though both clusters consisted mainly of smokers (Figure 2d). This was further supported by the identification of recently reported tobacco smoking-related signature SBS92 which we observed in the LX group alongside SBS4 (Figure 3b, 4, Supplemental Figure S7a,c). This finding is suggestive of selective, anatomical site-specific mutagenic effects of tobacco smoke, with distinct mutational processes operating in the smokers’ laryngeal tissues (SBS4, SBS92) and in the OC (SBS16).

### Tumors in the NIRF clusters are driven by endogenous mutagenic processes

NIRF-enriched clusters were characterized by increased levels of signature SBS1 in terms of both absolute and relative mutation counts (Figure 4 and Supplemental Figure S7a). Moreover, APOBEC signatures SBS2 and SBS13 were predominant in the APOBEC cluster (Figure 4 and Supplemental Figure S7a). Interestingly, in the APOBEC and 5mC deamin clusters, NIRF and drinker/smoker patients showed similar mutation spectra, except for the low presence of transcription strand biased T>C mutation pattern in the smoking/drinking patients’ tumors. The low presence of SBS16 signature in these patients suggests a weak contribution of alcohol/tobacco in the total mutation count in these clusters (Supplemental Figure S9). In sum, NIRF OC SCC are defined by elevated mutagenic activities of endogenous processes that result in the enriched presence of COSMIC signatures SBS1 and SBS2/13.

**Figure 4:**
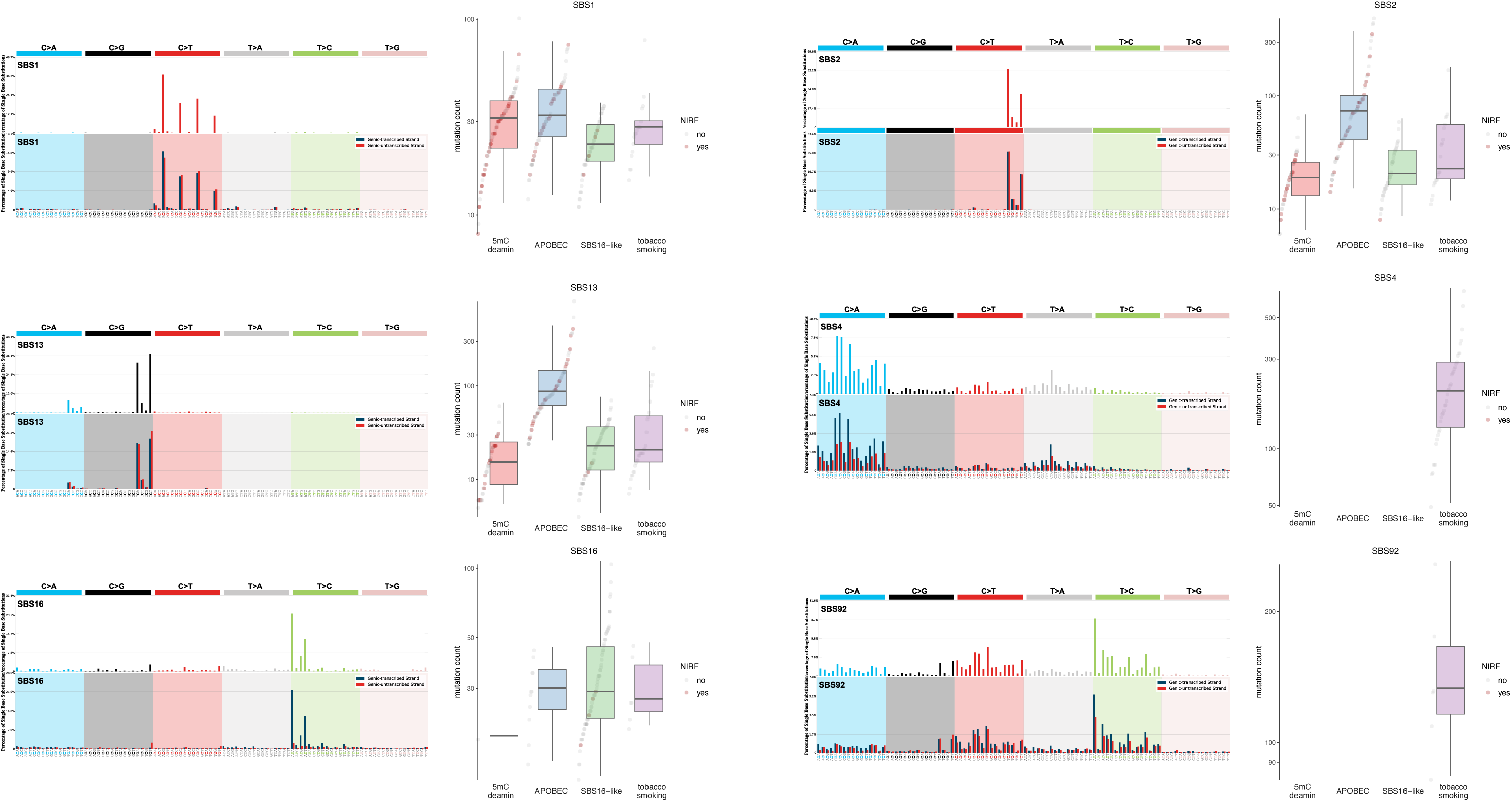
The main cluster-defining *de novo* signatures fitted by using MSA to their COSMIC signature contents. Top-left panels: the 96-channel COSMIC signatures; bottom-left panels: the corresponding transcriptional-strand asymmetry signature versions. The boxplots (right panels) show the per-cluster distribution of mutation counts attributed to each COSMIC signature.

### Cancer driver gene mutations in the NIRF-enriched clusters

The presence of distinct mutational processes predominantly operating in each cluster suggests potential differences in the natural histories of the disease development. To test this hypothesis, we determined mutated genes that were under selective pressure (see Methods). We identified 26 mostly recurrently mutated genes under positive selection with the following distribution: 5mC deamin cluster – 18/26, APOBEC cluster – 17/26, SBS16-like cluster – 11/26, tobacco smoking cluster – 6/26 (Figure 5a).

**Figure 5:**
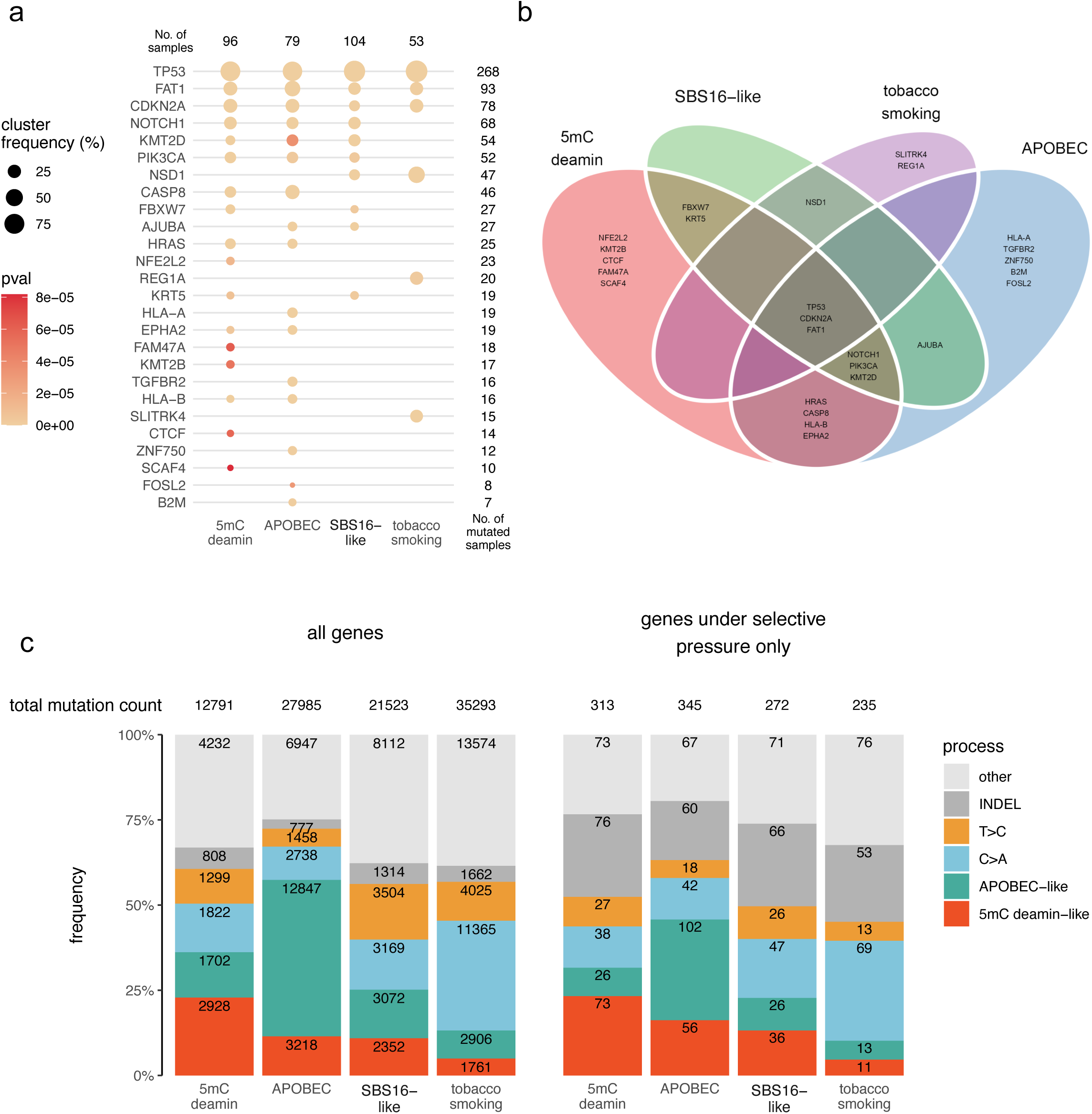
Cluster distribution of the identified non-silently mutated genes that are under selective pressure. (a) Summary of the mutated genes under selective pressure identified by the non-synonymous/synonymous mutation ratio approach (see Materials and Methods). The number of samples in each cluster is shown in the top row, and number of cases with a mutated gene (with symbols listed on the left) is shown on the right. P-values for substitutions were obtained by Likelihood-Ratio Tests as described ^67^ and cluster frequency indicate percentage of sample in each cluster mutated for a given gene. (b) Venn diagram showing the distribution of mutated genes under selective pressure across clusters. (c) Relative distribution of signature-matched mutation processes for all mutated genes in each cluster (left) and mutated genes that are under selective pressure (right). Numerical values in the graphs indicate the underlying mutation counts.

The mutated tumor suppressor genes *TP53*, *CDKN2A* and *FAT1* were selected for in all clusters, whereas various mutated epigenetic modifiers under positive selection exhibited more cluster specific distribution. Mutations in the histone methyltransferase *NSD1* were restricted to the tobacco smoking clusters (tobacco smoking and SBS16-like) while *KMT2B* mutants were only detected in the 5mc deamin cluster. Interestingly, genes involved in mitogen-activated protein kinase cascade components (*HRAS* and *EPHA2*) were mutated and selected for only in the NIRF-enriched clusters. The altered antigen presentation system (mutated *HLA-A*, *HLA-B, B2M genes*) and immune cytotoxic response (mutated *CASP8 gene*) were also specific to the NIRF-enriched clusters (APOBEC and 5mC deamin), suggesting the presence of immune pressure in these clusters (Figure 5b).

### The functional impact of mutational signatures

To address the impact of the key mutational signatures on all mutated genes, we selected the substitution sequence contexts that were most representative of a given mutagenic process and examined their presence in the mutated genes. The APOBEC (APOBEC-like) mutational process was represented by TCW, (where W = A or T), and the 5-methylcytosine spontaneous deamination was represented by XCG (X = either of A, T, C, G). As expected, mutations representing the APOBEC signature were more frequent in the APOBEC cluster while the 5mC deamin-like type mutations were also higher in APOBEC and 5mC deamin cluster. Furthermore, the C>A mutations (dominant in SBS4) were enriched in the tobacco smoking cluster and the T>C mutations in the SBS16-like cluster. Nearly identical distribution was observed for all mutated genes as well as the subset of genes under selective pressure (Figure 5c), suggesting that the overall processes mutagenizing single bases identified for each cluster were also affecting the genes selected for during cancer development. Furthermore, we observed that the selected genes were relatively more enriched for small insertions/deletions (indels) compared to all mutated genes (Figure 5c). Collectively, these data indicate the key endogenous mutational processes acting primarily on cytosine residues that characterize the overall as well as driver gene mutation landscapes in NIRF oral cancers.

### NIRF-enriched clusters exhibit keratinization and antimicrobial response gene expression programs

The causes of the SBS1 and SBS2/13 activity increase in the NIRF-enriched clusters are not known. To address this and to gain deeper functional and biological insights into the HNSCC patient stratification based on mutational signatures and the robustness of this approach, we performed DNA methylation analysis and differential gene expression and pathway analysis of the available TCGA methylome and transcriptomic data.

Previous studies have described distinct methylation profile in HNSCC subsites, including Brennan et *al.* who have reported five DNA methylation subtypes (Non-CIMP/CIMP Atypical, NSD1/ Stem-like Smoking and HPV+) using DNA methylation associated to mRNA expression (MethylMix). In addition to characterizing different epigenetic modifiers under selective pressure in the mutational signature-based clusters (Figure 5), we studied the distribution of the above methylation patterns in each cluster. Most patients in the two NIRF-enriched clusters harbored atypical, non-smoker DNA methylome patterns (Supplementary Figure S10).

Gene expression analysis of the NIRF-enriched clusters (see Methods) revealed higher expression of genes involved in the keratinization pathways (skin/epidermis development, keratinization, keratinocyte differentiation) (Figure 6a and b). In contrast, such activation of the keratinization pathway was absent in the tobacco smoking cluster (Supplemental Figure S11). Additionally, the NIRF-enriched clusters characteristically exhibited increased antimicrobial humoral response pathway activation (involving the genes *S100A7*, *KLK7*, *CXCL11*, *CST9*, *WFDC12*, and *DEFB4A*) (Figure 6a and b).

**Figure 6:**
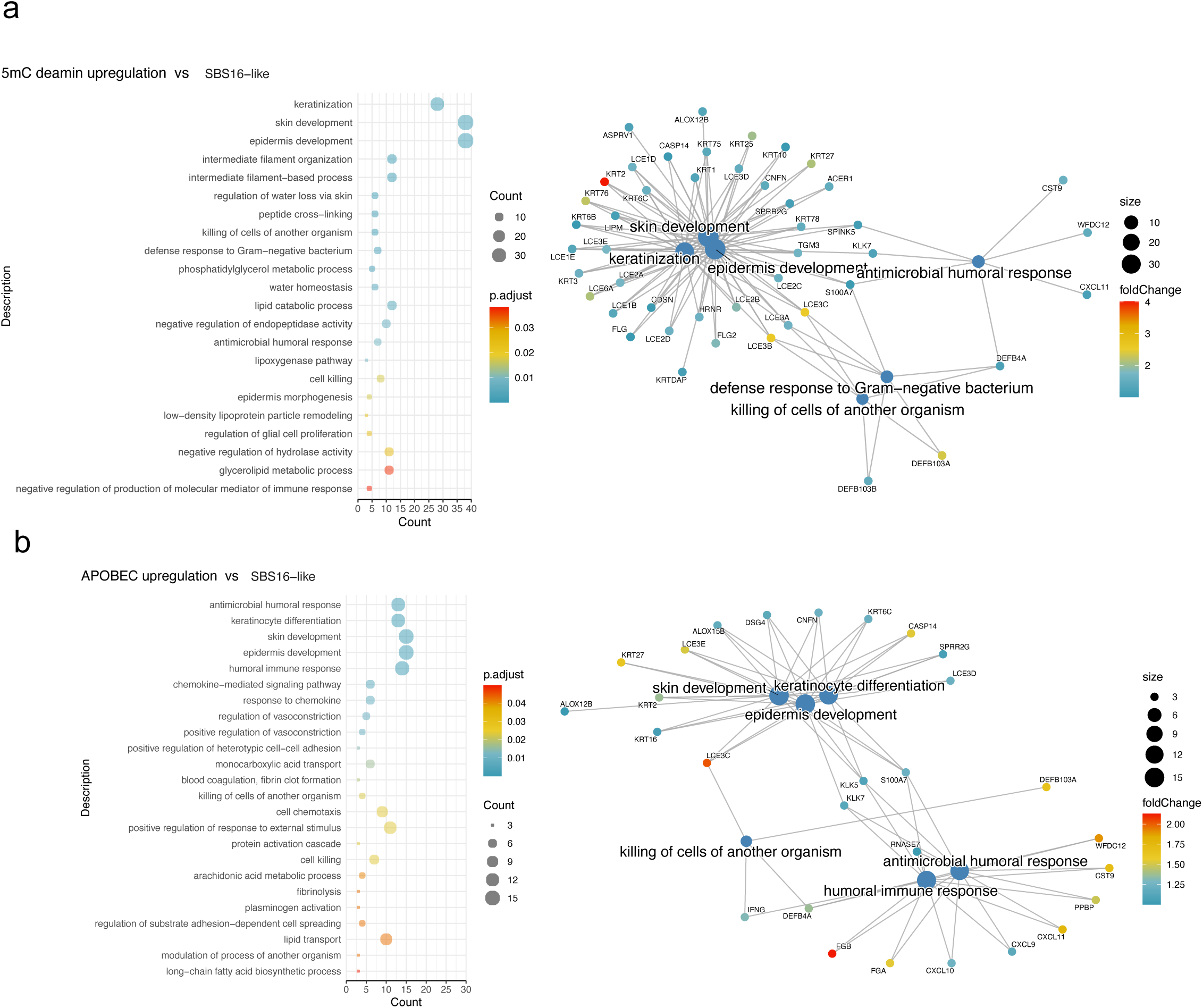
Enrichment of the clusters for functional biological processes. (a) Pathway enrichment for genes differentially expressed in the 5mC deamination cluster (a) or APOBEC cluster (b) using the SBS16-like cluster as reference. Pathways with enrichment p-values < 0.05 are listed on the left alongside count indicators representing the number of genes enriched in each pathway. Network diagram (right) shows selected top pathways of interest and their respective differentially upregulated.

We next investigated whether the activation of APOBEC enzyme activity revealed by the APOBEC-directed mutagenesis is associated with a specific immune status of patients in the NIRF-enriched clusters. APOBEC-induced mutations in cancer have been shown to be generated by the APOBEC3A and APOBEC3B enzyme, with distinct isoform-specific substrate preferences ^31^. APOBEC3A preferentially targets the YTCA sequence (Y = pyrimidine bases) whereas APOBEC3B is more prone to mutate the RTCA sequences (R = purine bases). We analyzed the APOBEC mutation context and observed a higher APOBEC3A activity marked by the enriched YTCA context in the APOBEC cluster (Figure 7a). The APOBEC3A mRNA expression was elevated in the APOBEC cluster indicating that APOBEC3A remained active at the time of tumor sampling (Figure 7b). Interestingly, the APOBEC3A expression in the APOBEC cluster correlated with activated genes and pathways involved in the bacteria-specific immune response (Figure 7c). In sum, the mutational signature-based patient clustering coupled with gene regulation/expression analysis suggest a possible etiology of the NIRF OCSCCs associated with cell keratinization and immune responses against bacterial insults.

**Figure 7:**
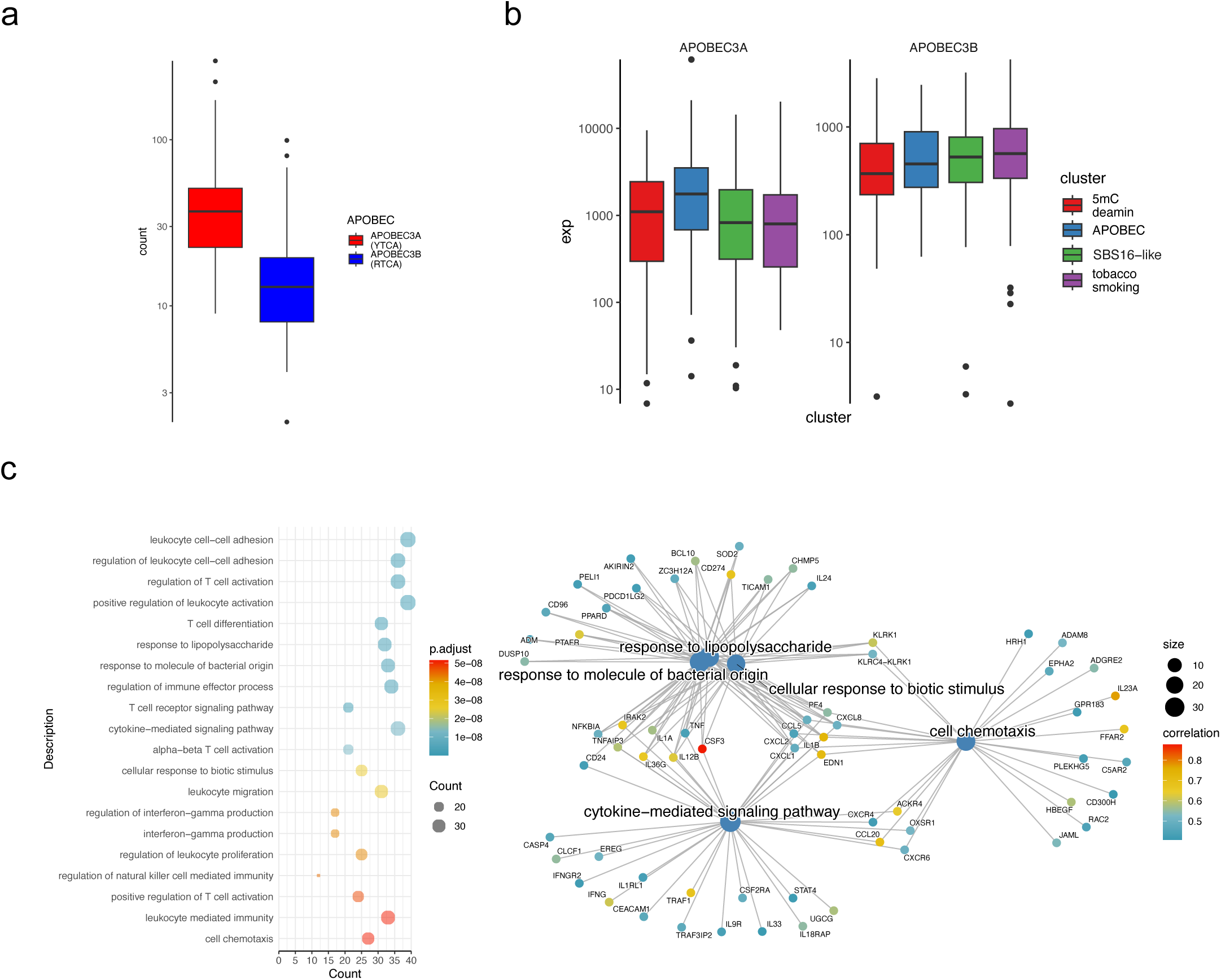
The origin of APOBEC mutagenesis. (a) mutation counts in the APOBEC cluster for APOBEC3A (targeting the YT<u>C</u>A context, where Y stands for pyrimidine bases) and APOBEC3B (RT<u>C</u>A context, R standing for purine bases). (b) The APOBEC3A and APOBEC3B mRNA expression across clusters. (c) Top 20 pathways enriched for genes with levels correlated with APOBEC3A expression.

## Discussion

Here we established that OC and LX SCC can be stratified according to anatomical subsite-specific mutational signatures and distinct sets of recurrently mutated, selected-for driver genes reflecting various histories of the cancer formation process. Importantly, such mutational signature-based patient stratification allows to discern the molecular landscapes of previously unrecognized cancer subtypes.

One example addressed here is the NIRF OC SCC, and specifically mobile tongue SCC in young patients, currently an insufficiently characterized clinical entity of increasing incidence. Our comprehensive analysis did not reveal mutational signatures suggestive of specific environmental mutagens for the NIRF-enriched patient subgroups. The NIRF OC SCC in the 5mc deamin and APOBEC clusters correlated with two endogenous mutational processes targeting cytosine residues. The 5mc deamin group was dominated by the activity of mutational signature SBS1, in keeping with a similar finding reported previously in another cohort ^18^. SBS1 is enriched for [C>T]pG mutations and it arises from endogenous processes, including spontaneous deamination of methylated cytosines, or enzymatic effects such as those exerted by the APOBEC cytidine deaminase family ^32–34^. The resulting T:G mismatch can be corrected by the DNA mismatch repair system before DNA replication. Consequently, the SBS1 mutation pattern can accumulate in the stem cells over the course of lifetime in relation with the frequency of their division, approximately reflecting the advancement of age ^26^.

While the NIRF-enriched clusters (5mc deamin and APOBEC) aligned with the expected NIRF clinical presentation, the enhanced SBS1 clock-like activity within the 5mC deamin cluster was somewhat surprising as patients in that group tend to be younger than the typical HNSCC patients. The SBS1 clock-like accumulation has also been described in differentiated cells such as neurons, suggesting that it can also be fixed in the genome in the absence of cell division, by an unknown process ^35^.

It remains unclear whether the two NIRF-enriched clusters represent distinct disease types or the same clinical entity with a subset marked by overactivation of the APOBEC3A enzyme and related mutagenesis. In support of the single clinical entity scenario, both NIRF-enriched clusters harbor higher SBS1 mutation counts, although the underlying mechanism for SBS1 elevation remains unclear. Moreover, the two NIRF-enriched clusters appeared to have acquired a specific immune escape response, as indicated by the positive selective pressure on mutated genes involved in Class I MHC antigen presentation system (*HLA-B*, *HLA-B, B2M*) and immune cytotoxic response (*CASP8*) suggesting the presence of a strong immune cytolytic control mechanism ^36^. The specific immune status of NIRF-enriched clusters is further underscored by differential gene expression profiles showing the presence of bacteria-specific immune responses in the NIRF-enriched clusters. Recent findings on intracellular bacteria invading the tumor cells offer an attractive putative explanation of such distinctive immune status ^37–40^. Our observations suggests that inhibitors targeting the PD1 axis will have limited effects on SCC exhibited by NIRF as defective antigen presentation is a source of primary or acquired resistance to immune checkpoint inhibitors ^41^.

It should be noted that while APOBEC activation in HNSCC has been previously linked to HPV infection ^28^, this study focused on subsites with low HPV prevalence, and the detection of HPV positivity in the APOBEC cluster was rare. Other viral origins of OC SCC in non-smoker and non-drinker patients had been proposed previously ^42^, warranting further investigations. In our study, APOBEC3A expression correlated with the antibacterial response which may represent an alternative point of departure for APOBEC activation. However, if the cancers in the 5mc deamin and APOBEC clusters are different manifestations with a common origin, it remains to be understood why patients in the APOBEC group, in addition to APOBEC activation, present with a distinct OC SCC localization and at an older age.

The signature SBS16 has been previously proposed to be linked to alcohol consumption. ^43–47^ Yet, it was also observed in tumor and non-tumor tissues of patients without previous history of alcohol exposure ^43,45–47^ and in HNSCC linked to tobacco smoking. Here we established that SBS16 mainly manifested in the OC tumors of smokers, especially those localized in the floor of the mouth. In keeping with previous studies showing that SBS16 can form in the absence of substantial alcohol exposure history ^48–52^, SBS16 was found in smokers’ SCC and at higher levels in SCC of smokers who were also drinkers. However, there were only 12 cases of non-smoking drinkers with OC and none harbored signature SBS16. This suggests that in HNSCC, signature SBS16 forms mainly due to the combined effects of tobacco smoking and alcohol drinking, or smoking only, rather than to alcohol exposure alone. This is in agreement with previous observations made in two independent cohorts, Plath *et al.* ^18^ and Gillison *et al.*^17^. Additional support for the association between tobacco smoking and SBS16 has been recently documented by a study on somatic mutagenesis in the non-cancerous bronchial epithelium ^53^.

Thus, alcohol and tobacco might elicit the same mutational process or synergize to drive mutagenesis manifesting primarily in the floor of the mouth. Alexandrov *et al.* reported that tobacco consumption can increase the rates of SBS5 mutagenesis in OC ^20^ yet given the similarity between SBS16, SBS5 and also SBS92 and the by-default inclusion of SBS5 in the SPE-based decomposition process, a role for overfitting of SBS5 is likely and should be considered. Indeed, we show that limiting the SBS5 overfitting favors the identification of SBS16 in the OC and makes SBS92 clearly discernible in tobacco smoking-associated laryngeal cancers. It should be noted that the analysis reported here is based on whole-exome sequencing with some samples harboring lower mutation counts, especially in the 5mC deamin and SBS16 clusters. Lower mutation counts can make the conditions for signature extraction less optimal, potentially affecting the diverse outcomes observed upon the SPE signature extraction and MSA signature assignment due to SBS5 overfitting.

We next observed that in tobacco smokers, the mutational processes and the overall mutation burdens differed between the OC and the LX, with the median mutation count nearly 10-fold higher in the LX. The mutational signatures SBS4 and (to a lesser extent) SBS92, both related to tobacco smoking were the main contributors to the mutational landscapes of the laryngeal tumors, yet they were barely detectable in the OC cancers of smokers.

Site-specific differences in mutation patterns in patients exposed to the same carcinogens might be due to the anatomical subsite susceptibility or protective capacity. For example, saliva has been shown to curb mutagen activity ^54–58^, a property that can explain the lower mutation burden observed in the OC. Moreover, mutagen concentration during the oral preparatory stage of swallowing ^59^, when the bolus is held in the anterior part of the floor of the mouth, could explain the higher proportion of SBS16 at that subsite. However, the precise molecular mechanisms underlying the signature SBS16 formation warrant further investigations.

Our study shows that OC and LX SCC follow distinct development trajectories, as indicated by the recurrently mutated genes under positive selection. While the selected-for mutations in the tumor suppressor genes *TP53*, *CDKN2A* and *FAT1* were observed in all patient-signature clusters, mutated epigenetic modifiers differed between the individual clusters. The recurrently mutated histone lysine methyltransferase *NSD1* was only detected in clusters enriched with smokers (SBS16-like and tobacco smoking) irrespective of the cancer subsite, suggesting that mutated *NSD1* could release a constraint imposed by tobacco exposure. This result corroborates several previously published observations, and it suggests potentially interesting clinical implications. Previous epigenomic analysis of the first set of 279 TCGA patients, detected a hypomethylated subgroup with loss of function of *NSD1* among 4 distinct methylation pattern (Hyper-methylated, Normal-like, Hypo-methylated, CpG island methylated) ^60^. Later, Papillon-Cavanagh *et al.* ^61^ re-analyzed the updated TCGA cohort with 528 HNSCC samples to classify HNSCC cases into five distinct subgroups: a subgroup harboring *NSD1* or histone *H3K36M* alterations, a subgroup positive for HPV and three less defined subgroups. By combining methylation state with gene expression data, Brennan *et al.* ^62^ uncovered two “atypical” subtypes with or without methylated CpG islands. The same approach was used to resolve 5 distinct methylation patterns in a pan-cancer study focusing on squamous cell carcinoma ^63^. The previously described hypermethylated HPV positive group was identified in both cervical and head and neck cancer sites while inactivating mutations in *NSD1* were linked to a hypomethylated group including, in addition to head and neck sites, tumors originating in the lung, cervix and esophagus.

*NSD1*-deficient tumors have been described as immune-cold tumors, with lymphocytes restricted to the stroma of the tumor borders, failing to infiltrate the tumor ^64^. *NSD1* inactivation drives tumor evasion ^64–66^ by reducing interferon-stimulated gene expression. Conversely, restoring methylation with the EZH2 histone-lysine N-methyltransferase inhibitor re-establishes immune response ^64^, offering a potential therapeutic strategy for patients with NSD1 inactivation mostly present in the tobacco cluster and also in the SB16-like cluster.

To conclude, the HNSCC patient stratification based on mutational processes is a valuable approach in addressing new angles of these cancers using relevant public repository data. This approach allowed us to better decipher the molecular bases of the HPV-negative HNSCCs, re-categorizing them from the “typical” tobacco related HNSCC to site-specific clinical entities with distinct cancer development paths, with some resulting subgroups manifesting with distinct immune evasion strategies. Our study also represents the first molecular and genomic characterization of the NIRF OCSCC patients and a first-of-its-kind basis for further studies of this emerging pathological entity with increasing global incidence.

## Methods

### Samples and clinical data

United States HNSCC cancer incidence data were collected from Global Cancer Observatory (GCO) and Surveillance, Epidemiology, and End Results (SEER) websites [2022/12/02]. TCGA Clinical data (528 patients) were downloaded from the Genomic Data Commons [GDC release v33.0 downloaded on 2022/05/11]. Clinical, demographic and lifestyle variables (cancer subsite, age, sex, tobacco smoking, alcohol consumption) for all available whole exome sequenced (WES) samples were extracted. After exclusion of files corresponding to hypopharynx and oropharynx the dataset included OC (n=314) and LX (n=111) SCC samples. Clinical and demographic variables extracted from TCGA were as described in Table 1. Cases were classified into three age groups (young ≤ 40 years, middle-aged 41-69 years and old ≥70 years). Tobacco-smoking status was classified as lifelong non-smoker (less than 100 cigarettes in the lifetime) or ever smoker. Alcohol consumption status was classified as non-drinker (less than 1 drink/day) or ever-drinker. HPV infection status was obtained through the study of Campbell et al. ^63^. Briefly, HPV status was assessed independently for all samples in two different centers by two different techniques (DNA sequencing and PathSeq algorithm and RNA-seq expression levels. Cases were classified as NIRF if they were non-smokers, non-drinkers, and HPV-negative.

### Data pre-processing

Controlled-access single nucleotide variant files of 425 whole exome sequenced cancers from the TCGA-HNSCC cohort analyzed in this study were obtained from Genomic Data Commons [GDC release v33.0 downloaded on 2022/05/11], for each variant caller used available (MuSE, MuTect2, VarScan2 and Pindel). Single nucleotide polymorphisms and reads that did not pass the variant caller’s filters were removed. Somatic mutation spectra of samples were generated using SigprofilerMatrixGenerator (v1.2) with its R wrapper, SigProfilerMatrixGeneratorR (v1.1) ^13^. Somatic mutations included single base substitutions (SBS), small insertions and deletions (indels or IDs).

### Identification of rare mutational spectra

Signature bleeding may occur in the process of signature extraction when signatures identified in a small, highly mutated sample subset are ascribed to all samples. In order to detect mutation spectra outliers and mitigate potential signature bleeding, we first identified samples with high mutation burden by looking at mutation count distribution (Supplementary Figure S2a). Next SBS96 mutation spectra of samples with more than 600 mutations were compared using cosine similarity. Hierarchical clustering of those samples (using 1-cosine as distance) showed presence of nine distinct mutation spectra (Supplementary Figure S2b). Six of these spectra, with a mean spectrum exhibiting potential matches with COSMIC SBS signatures, were found in fewer than 5 samples and thus categorized as rare (Supplementary Figure S2c). One spectrum, identified in two lip cancers sample and one OC cancer sample, exhibited similarities to the UV light signature (SBS7). Additional two spectra (present in three samples) resembled various combinations of SBS signatures of DNA mismatch repair deficiency (SBS15, SBS20 and SBS21). One spectrum (in one sample) manifested with a strong C>G pattern enrichment similar to the COSMIC SBS39 signature. Finally, one spectrum (in one sample) showed no resemblance to a COSMIC counterpart, and another (in one sample) was only observed as a result of MuTect2 variant calling and is thus a possible artifact.

### Extraction of *de novo* mutational signatures and sample clustering

After removing the atypical samples with high mutation count (see above), signature extraction was conducted on the 416 remaining samples using SigProfilerExtractorR (default parameters; v1.1.4) for each variant caller (MuSE, MuTect2, VarScan2) separately. While up to 10 *de novo* signatures were extracted in both the SBS96 and SBS384 catalogs, the optimal number of mutational signatures was assessed based on both the mean sample cosine and average stability, and set to 4 *de novo* signatures. Samples were finally clustered using K-means clustering algorithm into groups according to their signature relative contribution. Using the 384-channel signature dimension increased the stability across variant callers (Supplementary Figure S4a), by reducing the number of patients switching between clusters, from 21% (88/416) to 16% (67/416). Consequently, we consistently used the 384 channel-based signature analysis throughout the study.

A heatmap of SBS384 signature extraction was generated for each variant callers (Supplementary Figure S4b) and sample content in each cluster was compared among the variant caller-specific results, using Venn diagram (Supplementary Figure S4c). For each cluster, samples at the intersection of the 3 different variants were considered to be the most stable and were used for the final clustering. This final sample (n=347) assignment to each cluster was used in the remaining analysis steps.

### COSMIC signature fitting

The all-COSMIC signature fitting by non-negative least squares (NNLS) was performed using MuTect2 variant caller-detected mutations, in samples with consistent clustering performance (see above). First, all COSMIC signatures were fitted to *de novo* extracted signatures using SigProfilerExtractorR (v1.1.4), using default parameters. Next, the MSA tool (v2.0) ^27^ was used to fit the COSMIC signature set proposed by SigProfilerExtractor or to the well curated catalogue of known COSMIC signatures (version 3.2) to each sample individually. As no SBS384 version of COSMIC signature was available at the time of the analysis, the SBS288 transcription strand bias versions of COSMIC signatures was used in each case, with optimal threshold suggested by the MSA tool. Signature relative contribution was used to assess clustering stability. For final signature attribution, signature activities that included zero value inside the 95% confidence intervals were pruned to prevent overfitting.

### Mutated genes under selective pressure

Genes under selective pressure were identified by computing non-synonymous to synonymous mutations ratio using dNdScv tool (v0.0.1.0) without covariate ^67^. A dedicated mutation set was created for this analysis using Pindel in addition to the other variant callers used (MuSE, MuTect2, VarScan2). Only SBS mutations or indels identified by at least two variant callers were retained. A mean VAF value was calculated for each resulting mutation. TCW (with W standing for A or T) was used as template to identify APOBEC-like mutations and XCG (with X standing for A,T,C,G) for 5mC deamin-like mutation.

### Differential Gene expression and methylation analysis

For gene expression analysis, open access RNA-Seq raw counts were download from GDC [GDC release v33.0 downloaded on 2022/05/11]. The analysis was focused on primary tumors exhibiting consistent clustering performance, resulting in 342 samples (see Extraction of *de novo* mutational signatures and sample clustering section).

Normalization and differential gene expression analysis of NIRF-enriched clusters (5mc deamin and APOBEC) were carried out against the other OC SBS16-like cluster using DESeq2 (version 1.38.3). As a control, the comparison of the tobacco smoking cluster to the SBS16-like cluster was performed.

Gene Ontology (GO) term enrichment analysis was conducted using the clusterProfiler package ^68^ (version 4.6.2), on differentially expressed genes (DEG) with adjusted p-values less than 0.05 and a log fold change greater than 1.

Methylmix subtype were derived from conducted by Brennan et all. ^62^. The NSD1 mutation status was determined using Ensembl VEP annotation extracted from single nucleotide variant files. Only mutations with a high impact score were considered as inactivating mutations.

## Data Availability

All data produced in the present study are available upon reasonable request to the authors

## Acknowledgments

This study was funded in part by the NIH/NIAAA grant R01AA029736 (J.Z.).

## Author contributions

The authors confirm contribution to the paper as follows: study conception and design: SD, BF, JZ, FV; analysis and interpretation of results: SD, BF, SS, LB, OP JZ, BC, FV; draft manuscript preparation: SD, BF, SS, LB, OP, BC, LL, IC, TR, JZ, FV. All authors reviewed the results and approved the final version of the manuscript.

## Disclaimer

Where authors are identified as personnel of the International Agency for Research on Cancer/World Health Organization, the authors alone are responsible for the views expressed in this article and they do not necessarily represent the decisions, policy or views of the International Agency for Research on Cancer /World Health Organization.

## Materials and Correspondence

(FV), or (JZ)

