## Supplemental Figures and Table for "Molecular landscapes of oral cancers of unknown etiology"

### Suppl. Figure S1

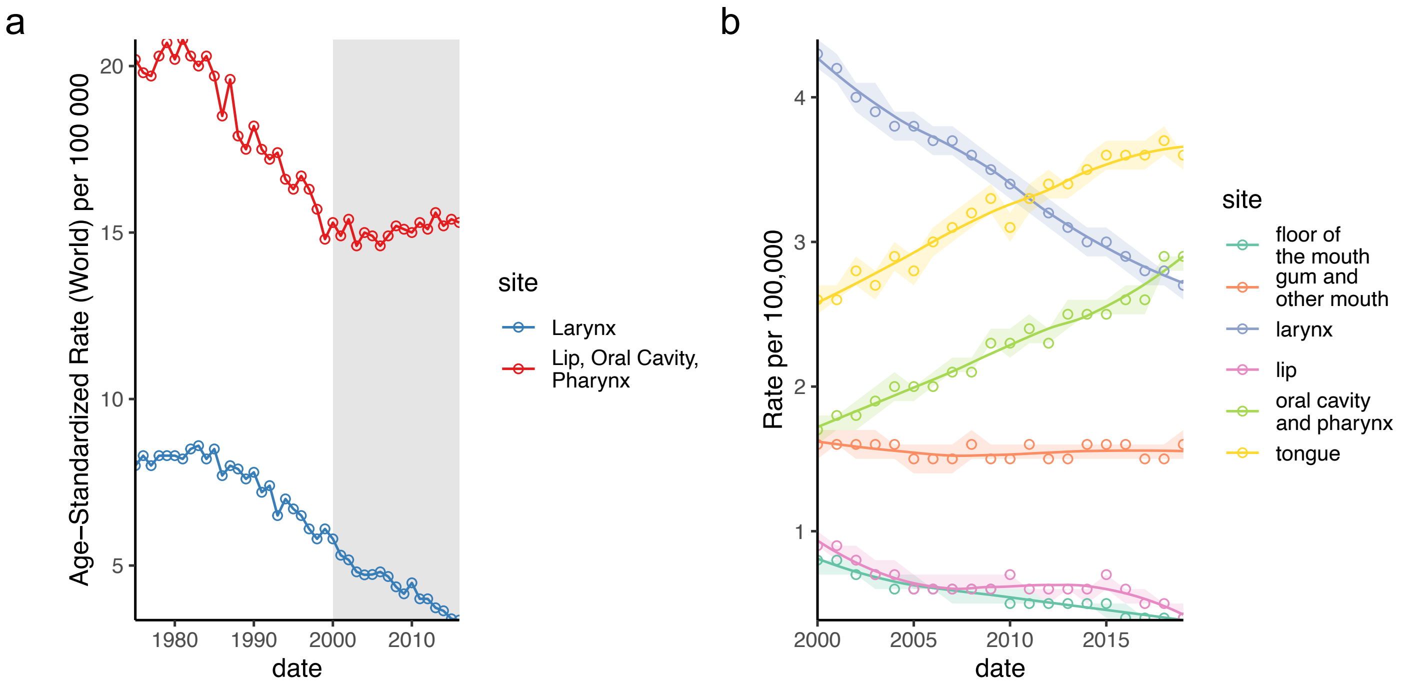


**Supplemental Figure S1:** Head-and-neck cancer incidence trends in the US. (a) Change in the worldwide age-standardized incidence rates of non-laryngeal head and neck cancers. Source: Global Cancer Observatory, IARC, WHO. (b) Site-specific incidence trends indicate marked increases for HPV-associated oropharyngeal and tonsillar cancers, and cancers of the mobile tongue. Source: SEER, NIH NCI.

### Suppl. Figure S2

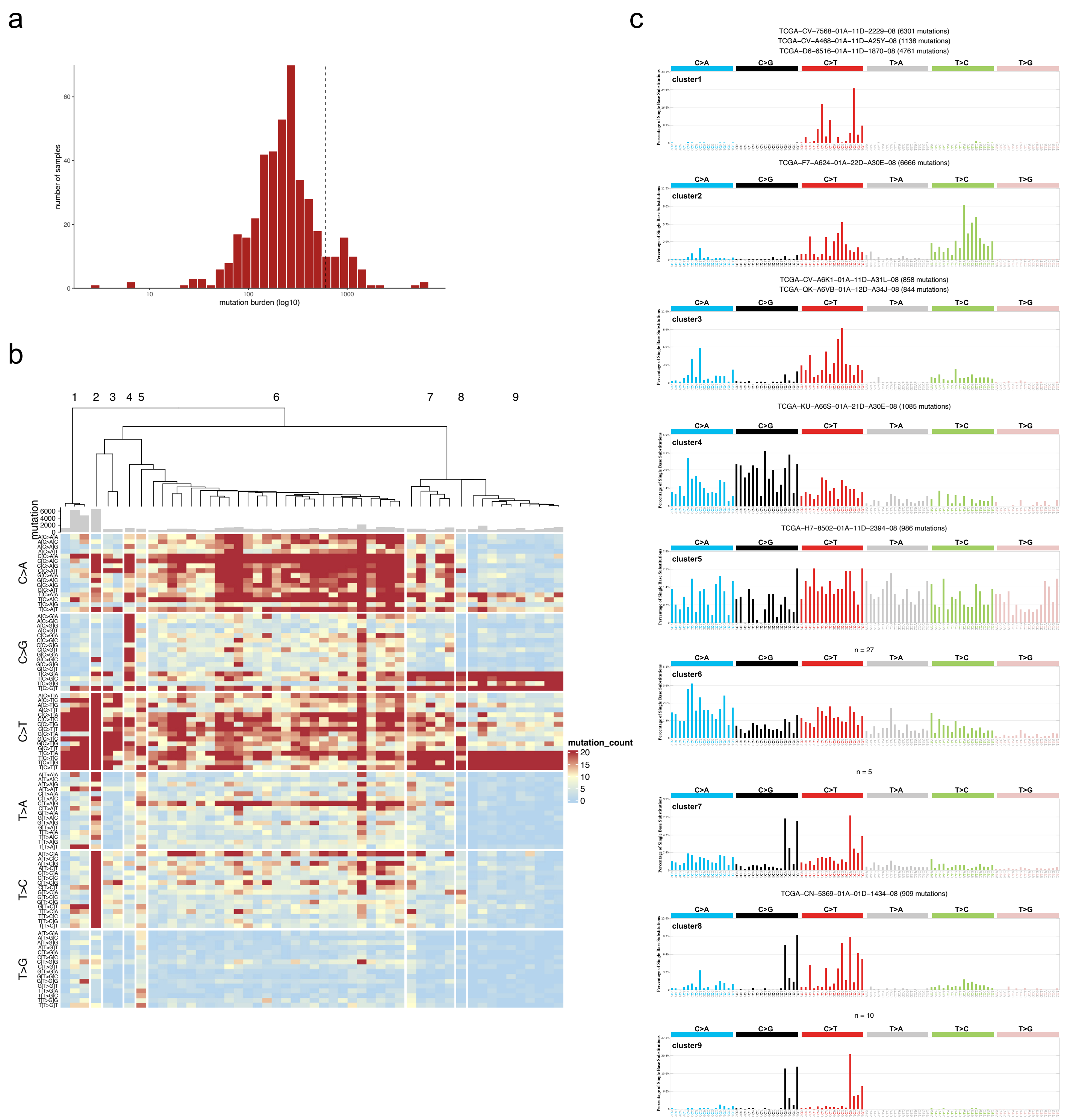


**Supplemental Figure S2:** Identification of rare mutational spectra. (a) Mutation burden distribution. (b) Heatmap representing mutation count in each SBS96 channel (row) for samples (column) with more than 600 mutations. Samples are ranked using hierarchical clustering with 1 - cosine as distance. Tree has been cut to create 9 clusters. (c) Mean mutation spectra for each cluster. For clusters with less than 5 samples, both sample name and mutation count are indicated, otherwise only number of samples is given.

### Suppl. Figure S3

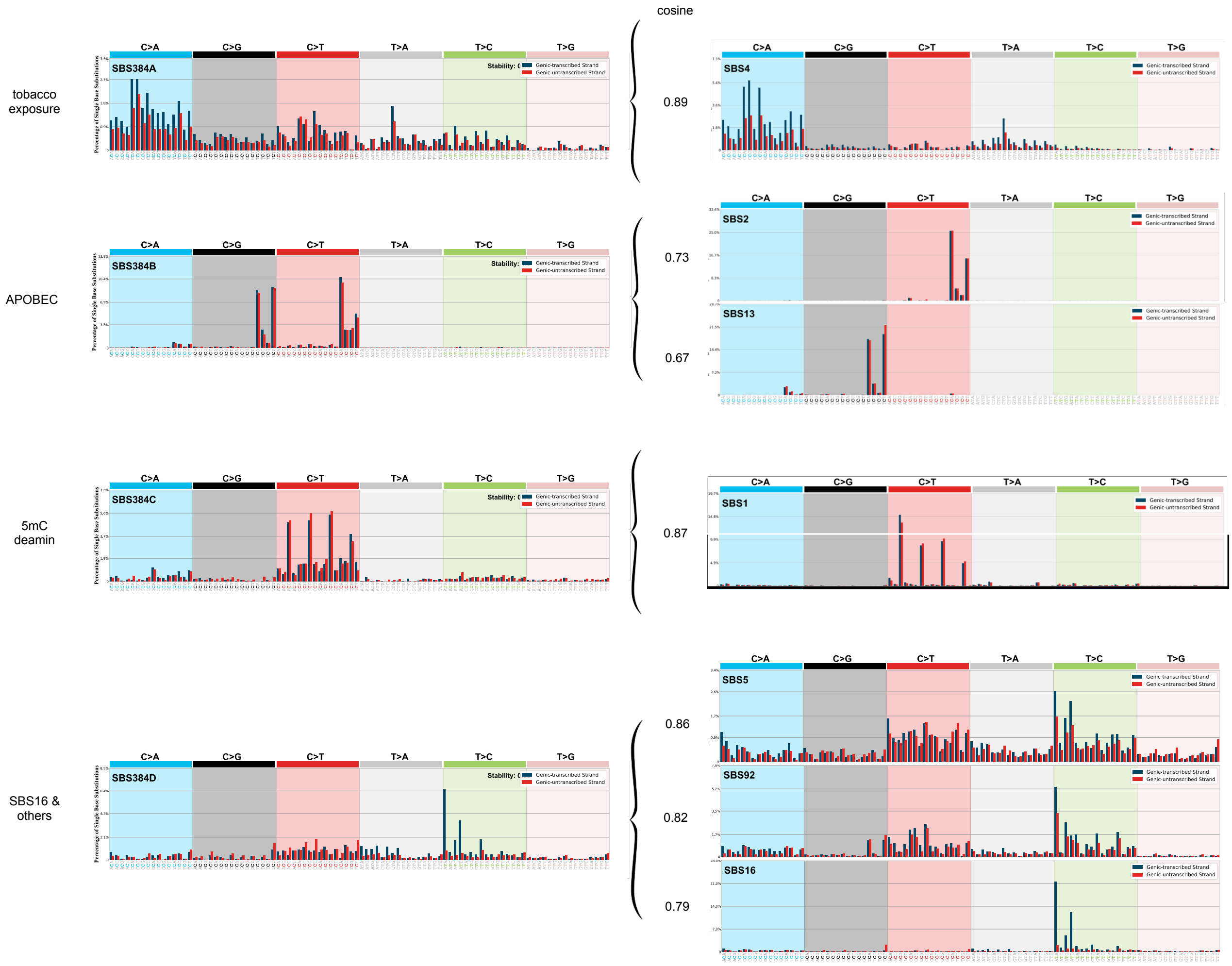


**Supplemental Figure S3:** *De novo* signatures identified after NMF extraction (A-D) with their putative COSMIC signature equivalents.

#### Suppl. Figure S4

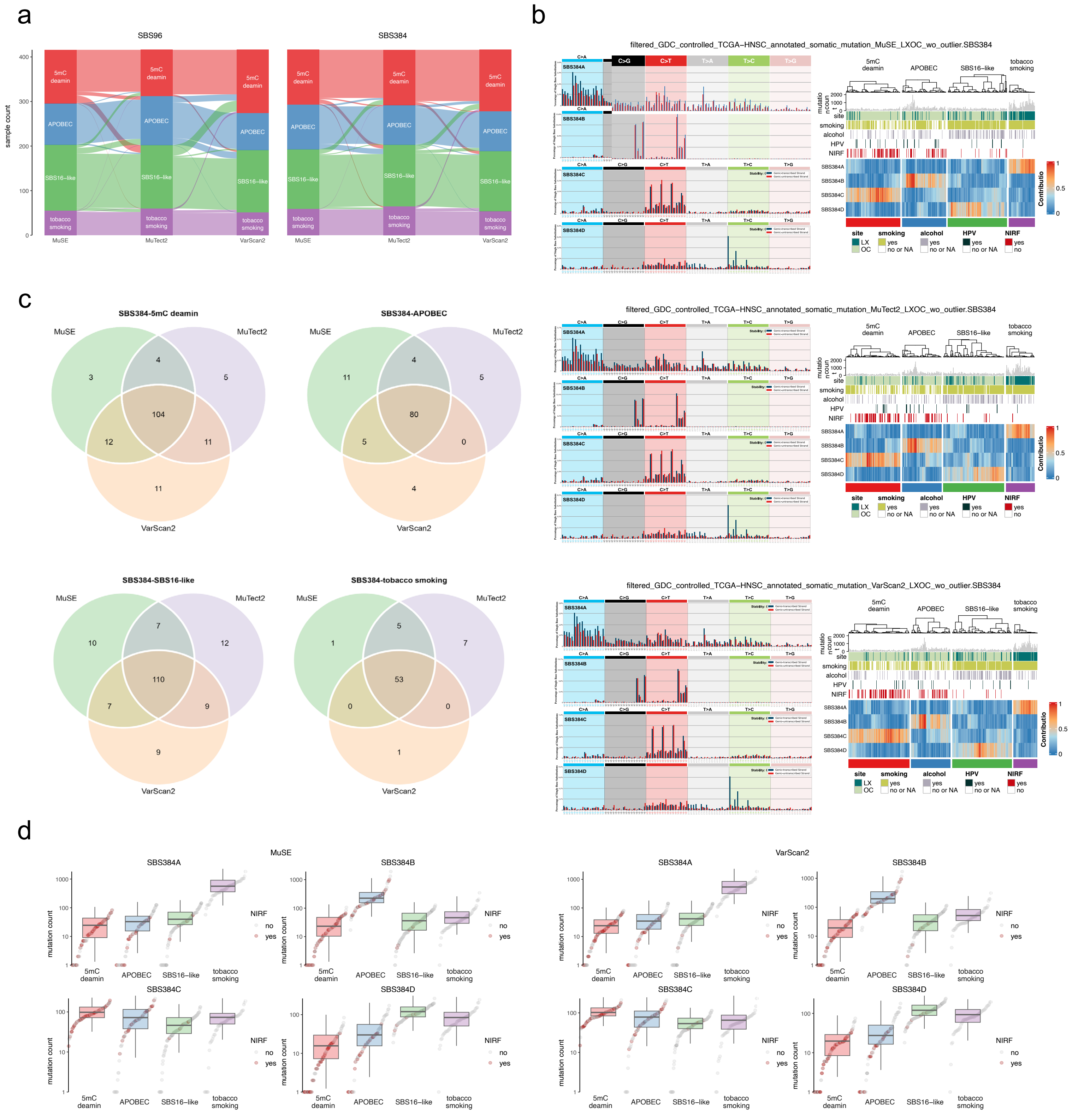


**SupplementalFigure S4:** Cluster stability for different variant callers or mutation spectrum dimensions. (a) Sample flow between clusters depending on the variant caller and the mutation spectrum dimension (SBS96- or SBS384-channel). (b) Mutation spectra and sample clustering for Muse, MuTect2 and VarScan2 variant callers at the SBS384-channel level. (c) Sample and cluster based Venn diagrams for MuTect2, VarScan2 and Muse variant caller-based SBS384-channel spectra. For the rest of the study, clusters included samples from the intersection of all variant callers. (d) Signature mutation burden for each cluster depending on each variant caller.

### Suppl. Figure S5

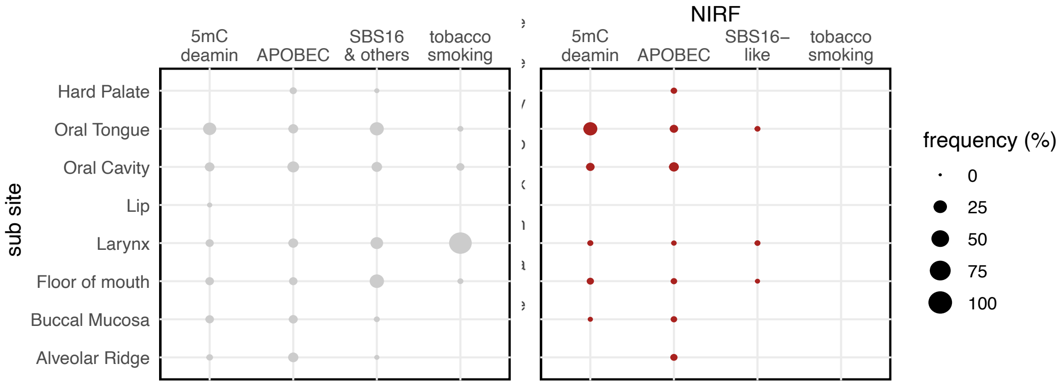


**Supplemental Figure S5:** Bubble plot showing the anatomical sub-site distribution of the NIRF and non-NIRF patient subsets in each cluster

### Suppl. Figure S6

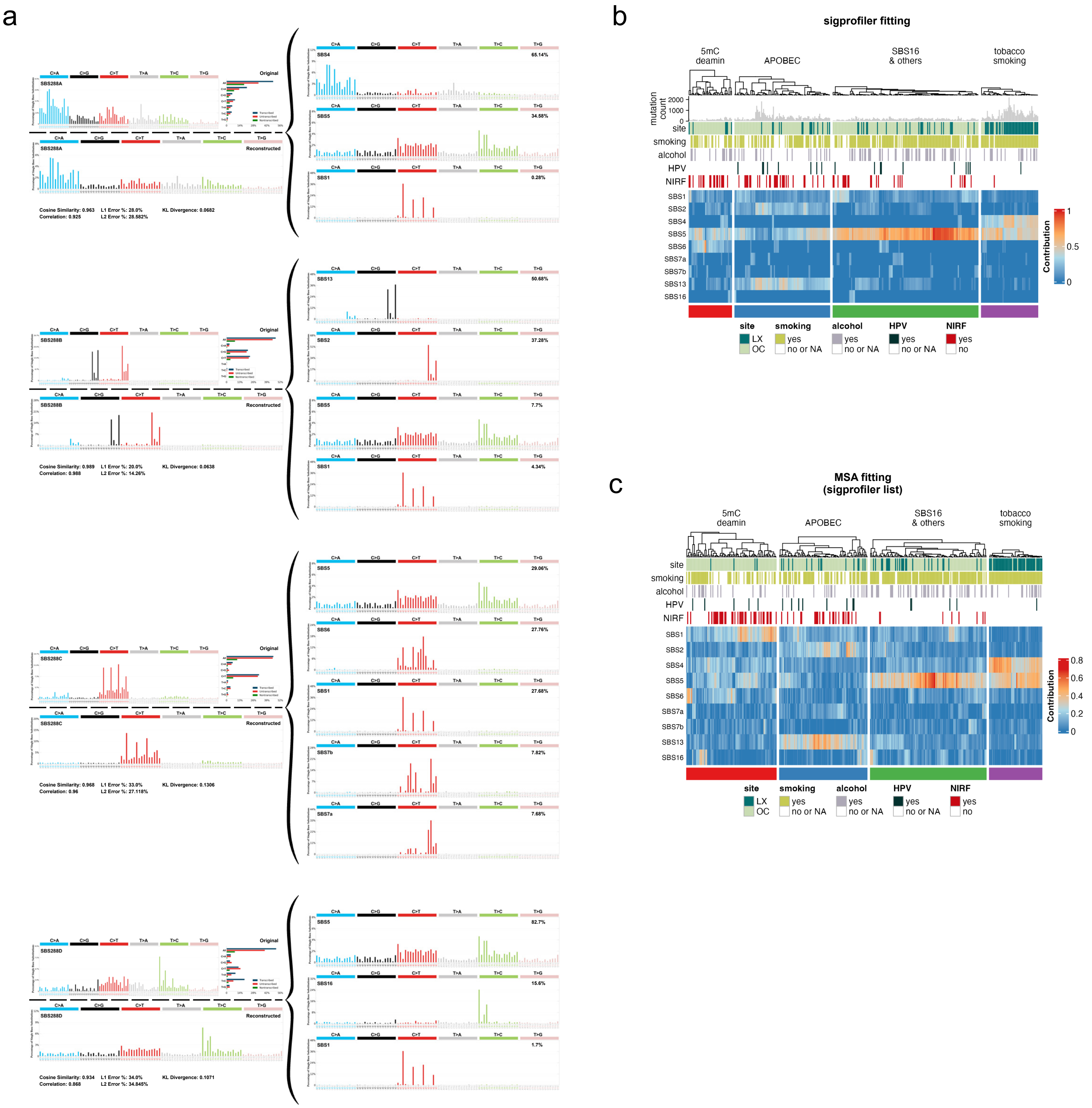


**Supplemental Figure S6:** SigProfiler-based fitting. (a) *De novo* signatures decomposed into COSMIC signatures by SigProfiler. (b) Heatmap and clustering based on the SigProfiler signature attribution. (c) Heatmap and clustering based on the MSA signature attribution using signature set proposed by the SigProfiler fitting.

### Suppl. Figure S7

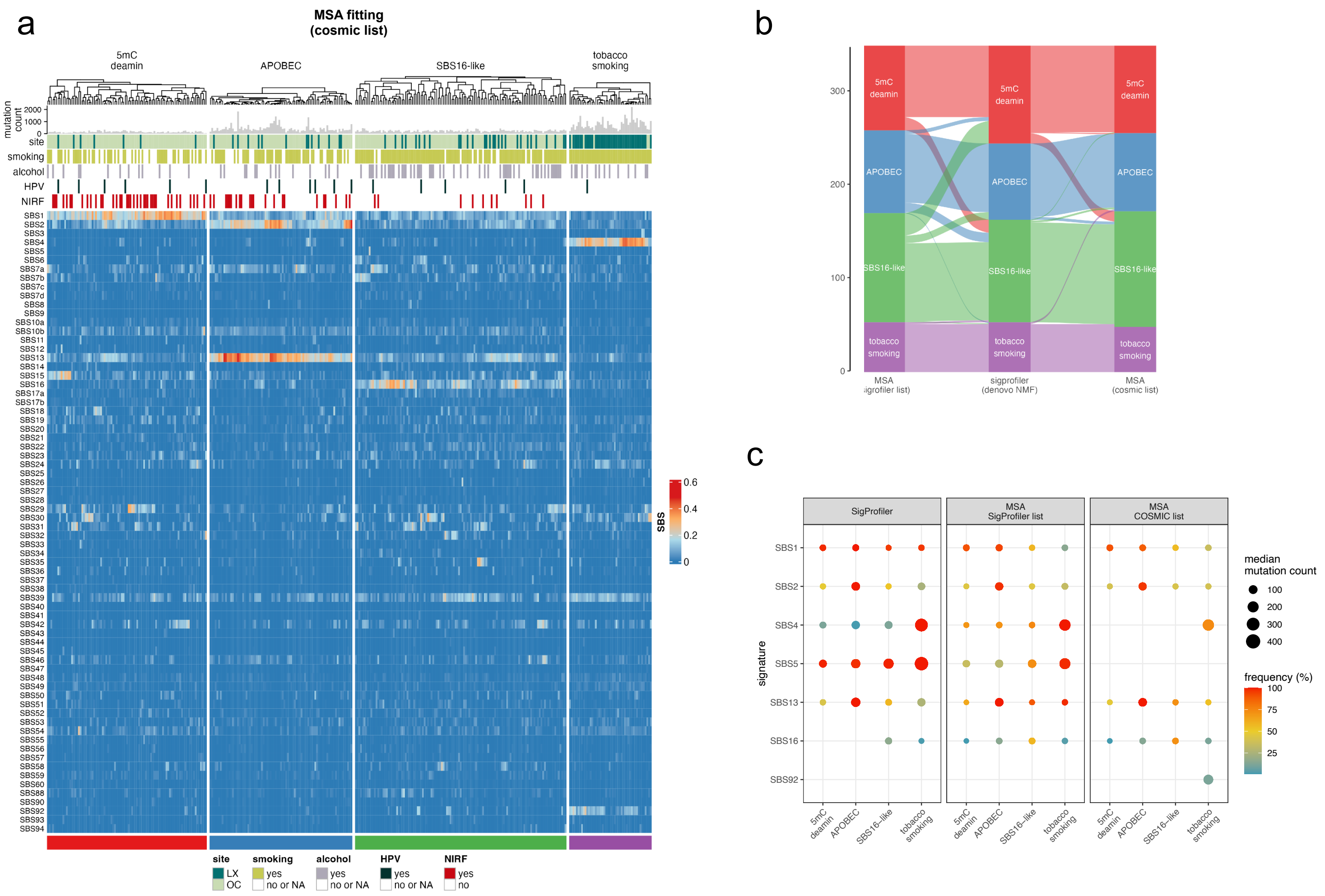


**Supplemental Figure S7:** A comparison of signature attribution by MSA or SigProfiler. (a) Heatmap and clustering resulting from MSA signature attribution using the whole COSMIC signature set. (b) Sample flow between clusters depending on the signature fitting strategy. (c) Bubble plot showing expected and discovered signatures, with median mutation count (size) and positive sample frequency (color) shown for each cluster and for the respective fitting strategies.

### Suppl. Figure S8

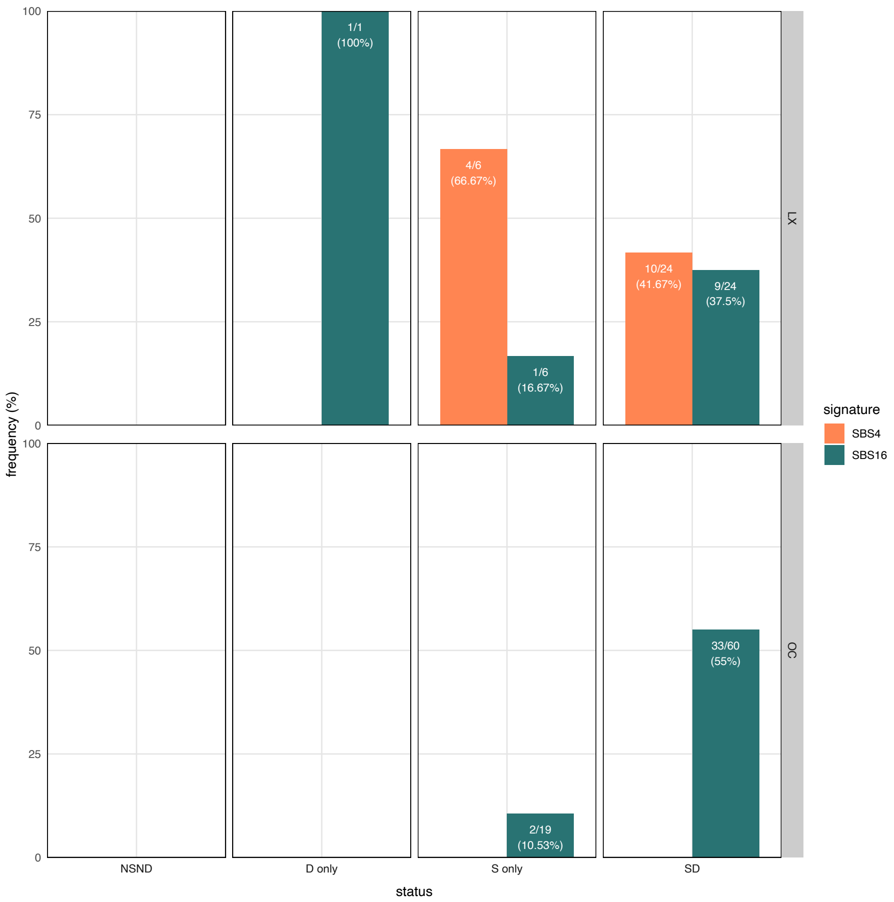


**Supplemental Figure S8:** Relative distribution of samples positive for signature SBS4 and/or SBS16, according to the anatomical site, smoking and alcohol consumption status (non-smoker non-drinker (NSND), drinker only (D) , smoker only (S), smoker drinker (SD))

### Suppl. Figure S9

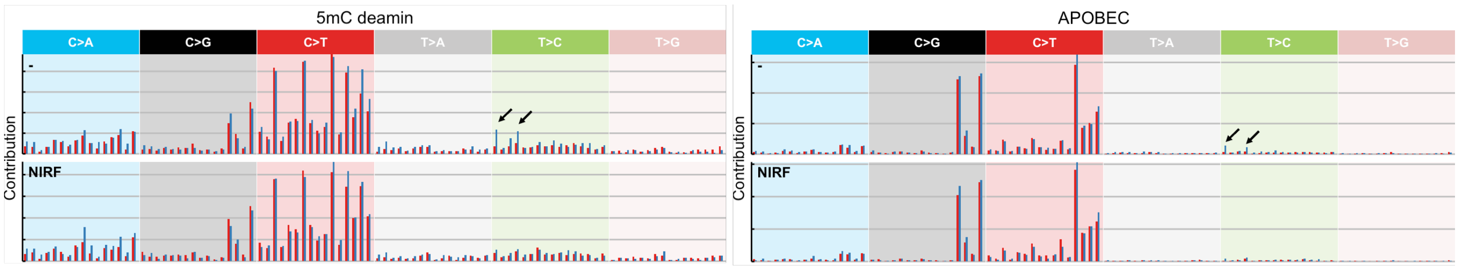


**Supplemental Figure S9:** Mean profile spectra of the NIRF and non-NIRF patients in the NIRF-enriched clusters (5mC deamin and APOBEC)

#### Suppl. Figure S10

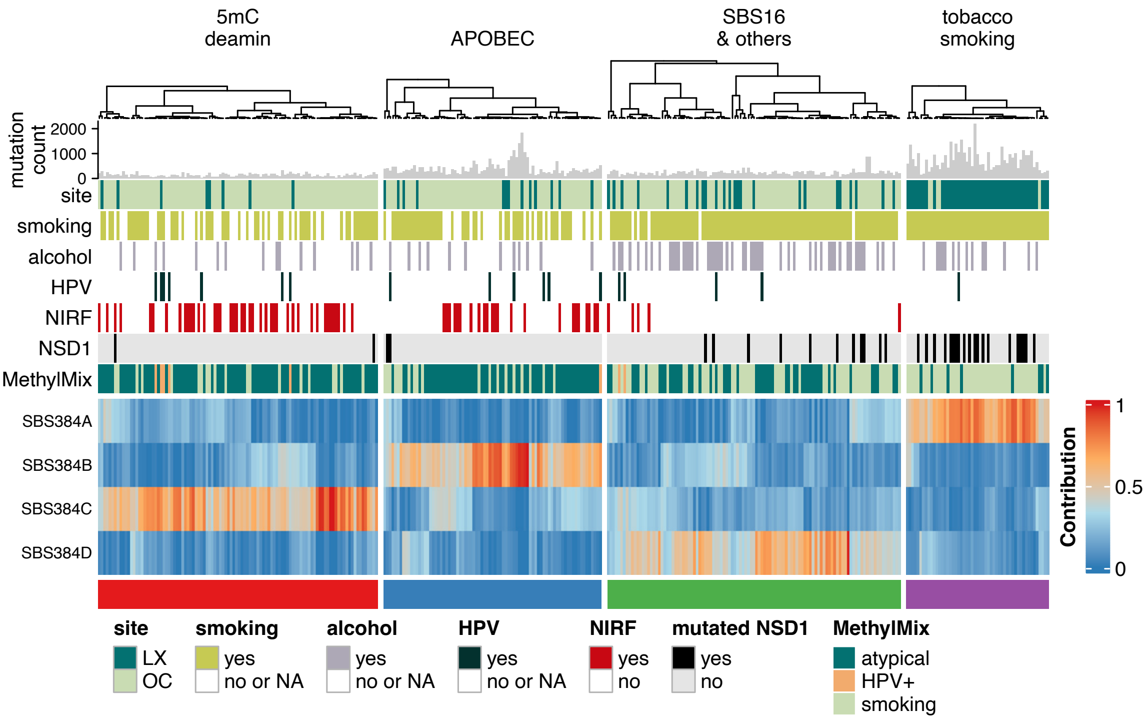


**Supplemental Figure S10:** Cluster methylation profiles. The cluster heatmap with the additional track showing the Methylmix DNA methylation subtypes, previously described by Brennan et al.[62] (atypical = Non-CIMP and CIMP subtypes, smoking = NSD1 and Stem-like subtype).

### Suppl. Figure S11

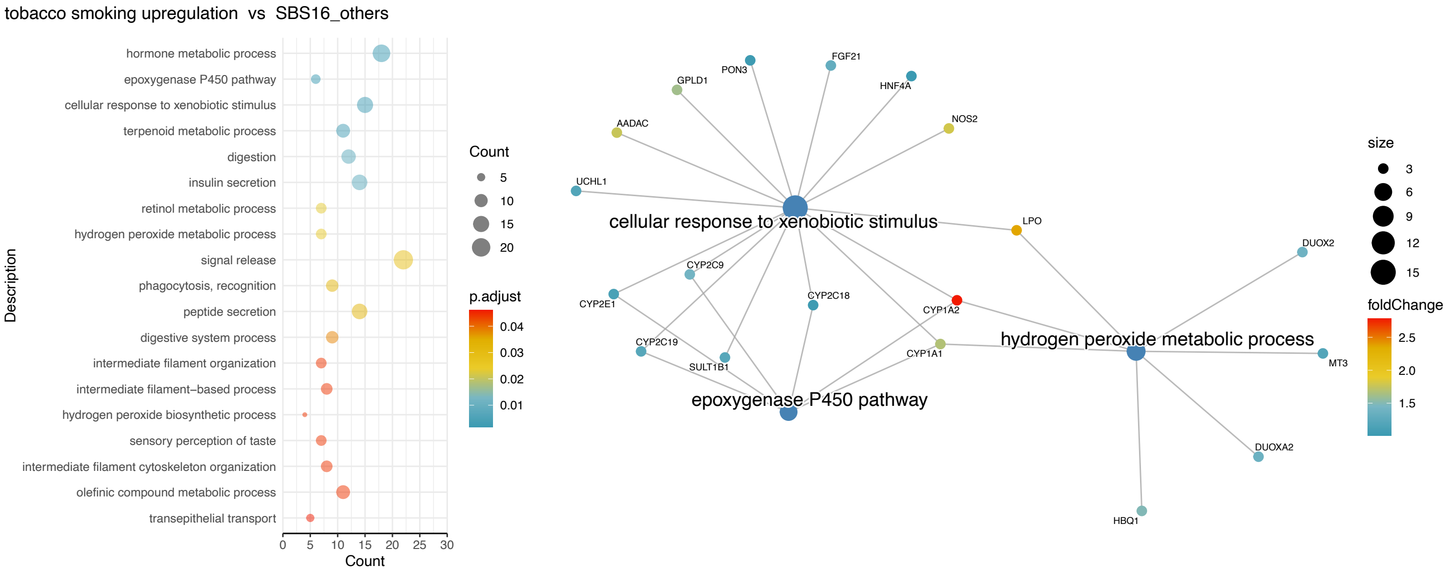


**Supplementary figure S11:** The functional enrichment of the tobacco-smoking cluster. Pathway enrichment for genes di!erentially expressed in the tobacco-smoking cluster using the SBS16-like cluster as reference. Pathways with enrichment p-values < 0.05 are listed on the left alongside count indicators representing the number of genes enriched in each pathway. Network diagram (right) shows selected top pathways of interest and their respective differentially upregulated.

### Suppl. Table S1

| | | S | | | NS | | | unknown | | |
|---|---|---|---|---|---|---|---|---|---|---|
| site | HPV | D | ND | unknown | D | ND | unknown | ND | unknown | Total |
| OC | - | 72 (55) | 19 (18) | 111 (87) | 8 (5) | 18 (16) | 52 (44) | 0 (0) | 6 (5) | 286 (230) |
| | + | 4 (4) | 0 (0) | 8 (5) | 2 (2) | 1 (1) | 3 (3) | 0 (0) | 0 (0) | 18 (15) |
| | unknown | 2 (1) | 1 (1) | 3 (3) | 0 (0) | 2 (1) | 1 (1) | 1 (1) | 0 (0) | 10 (8) |
| LX | - | 29 (22) | 7 (6) | 62 (54) | 1 (1) | 0 (0) | 5 (4) | 0 (0) | 3 (3) | 107 (90) |
| | + | 2 (2) | 0 (0) | 1 (1) | 0 (0) | 0 (0) | 0 (0) | 0 (0) | 0 (0) | 3 (3) |
| | unknown | 0 (0) | 0 (0) | 1 (1) | 0 (0) | 0 (0) | 0 (0) | 0 (0) | 0 (0) | 1 (1) |
| | subtotal | 109 (84) | 27 (25) | 186 (151) | 11 (8) | 21 (18) | 61 (52) | 1 (1) | 9 (8) | 425 (347) |

Initial dataset (Stable dataset)

**Supplementary Table 1:** Distribution of samples based on the anatomical site and exposure status, and the selection of a core sample subset upon exclusion of samples wih unstable behavior during data preprocessing (see Methods).
